# Neural Correlates of Remotely versus In-person Administered Symbol Digit Modalities Test in Multiple Sclerosis

**DOI:** 10.1101/2022.06.22.22276749

**Authors:** Korhan Buyukturkoglu, Jordan D. Dworkin, Victor Leiva, Frank A. Provenzano, Pamela Guevara, Philip L. De Jager, Victoria M. Leavitt, Claire S. Riley

## Abstract

**Background:** Prior studies in multiple sclerosis (MS) support reliability of telehealth-delivered cognitive batteries, although, to date, none have reported predictive external validity, i.e., relationships of cognitive test performance to neural correlates across administration modalities. In this study we aimed to compare brain-behavior relationships, using the Symbol Digit Modalities Test (SDMT), the most reliable and sensitive cognitive measure in MS, measured from patients seen via telehealth versus in-person.

**Methods:** SDMT was administered to individuals with MS either in-person (N=60, mean age=39.7) or remotely via video conference (N=51, mean age=47.4). Magnetic resonance imaging (MRI) data was collected in 3-Tesla scanners. Using 3D-T1 images cerebral, cortical, deep gray, cerebral white matter and thalamic nuclei volumes were calculated. Using a meta-analysis approach with an interaction term for participant group, individual regression models were run for each MRI measure having SDMT as the outcome variable in each model. In addition, the correlation and average difference between In-person and Remote group effect sizes across the MRI measures were calculated. Finally, for each MRI variable I^2^ score was quantified to test the heterogeneity between the groups.

**Results:** Administration modality did not affect the association of SDMT with MRI measures. Brain tissue volumes showing high associations with the SDMT in one group also showed high associations in the other (*r* = 0.83; 95% CI = [0.07, 0.86]). The average difference between the In-person and the Remote group effect sizes was not significant (β_Remote_ - β_In-person_ = 0.14, 95% CI = [-0.04, 0.34]). Across MRI measures, the average I^2^ value was 14%, reflecting very little heterogeneity in the relationship of SDMT to brain volume.

**Conclusion:** We found consistent relationships to neural correlates across in-person and remote SDMT administration modalities. Hence, our study can be considered a step towards providing predictive external validity to remote administration of the SDMT in MS.

**Highlights:**

1. Symbol Digit Modalities Test (SDMT) was administered to individuals with MS either in-person or remotely via video conference.
2. Administration modality did not affect the association of SDMT with MRI measures.
3. Brain tissue volumes showing high associations with the SDMT in one group also showed high associations in the other.
4. The average difference between the In-person and the Remote group effect sizes was not significant.
5. This study can be considered a step towards providing predictive external validity to remote administration of the SDMT in MS.

## 1. Introduction

Cognitive impairment is frequent and negatively impacts quality of life in multiple sclerosis (MS),^1^ and, therefore, it should be carefully evaluated and monitored.^2^ Traditionally this is done with in-person visits using comprehensive batteries of neuropsychological tests.^3–6^ However, during the COVID-19 pandemic, many MS patients deferred non-emergent outpatient appointments to avoid potential exposure.^7^ As a result, the field of neuropsychology accelerated the trend of evaluating and adopting test batteries that are deployed remotely to enhance patient safety.

Previous studies have reported concordant results between in-person and remote administration of cognitive tests in MS,^7–11^ supporting the reliability of remotely-administered cognitive batteries. Some of these studies focused exclusively on Symbol Digit Modalities Test (SDMT),^12^ which is generally considered to be the most sensitive cognitive measure for use in MS and is the most-utilized cognitive measure in MS clinical trials.^13–16^ Two recent studies showed reliability of remote administration of SDMT using a within-subject design.^7,9^ Here, to complement these earlier studies which focused on evaluating the reliability of the new video-conference based testing modality to standard testing, we undertook an effort to assess its practical use: the ability to use remote testing in a clinical research setting to replicate known, robust associations. This will provide predictive validity for remote administration of SDMT in MS.

SDMT is strongly correlated with gray matter volume in MS,^17–24^ however white matter microstructural damage and volume loss is also associated with cognition in MS.^25^ Therefore, we calculated global and local gray and white matter volumes (e.g., total cortical, deep gray matter, anterior thalamic nuclei, cerebral white matter volume) and hypothesized that SDMT, would show consistent relationships to these MRI measures irrespective of the modality of administration. Specifically, we compared the statistics derived from in-person SDMT measures paired with a research MRI to those derived from SDMT measures collected by video conferencing paired with the closest available clinical MRI.

## 2. Materials and Methods

The institutional review board of the Columbia University Irving Medical Center (CUIMC) approved the study protocols, and all participants provided written informed consent.

A total of 115 relapsing remitting (RR) MS patients were included in this study. Participants in the remote group were selected either from an ongoing study (MS Snapshot, a longitudinal cohort study recruited from the patient population at the Columbia MS Center (mean age=46.7, N=42) or from a pool of patients who were referred by a neurologist in our department for a neuropsychological evaluation (mean age=36.3, N=13) had their visits via telehealth between 2020 and 2022. For both groups, SDMT (oral version) was administered remotely via secure Zoom link in their home settings.

Participants in the in-person group were selected from an ongoing study cohort (MemConnect, mean age=39.7, N=60).^26^ For those participants, SDMT was administered at Columbia University Irving Medical Center during a research visit.

Following exclusion of four participants for whom brain tissue segmentation failed, analyses were performed on the remaining 111 participants (51 Remote, 60 In-person).

Demographic and clinical characteristics of the participants are presented in **Table 1**.

**Table 1.**
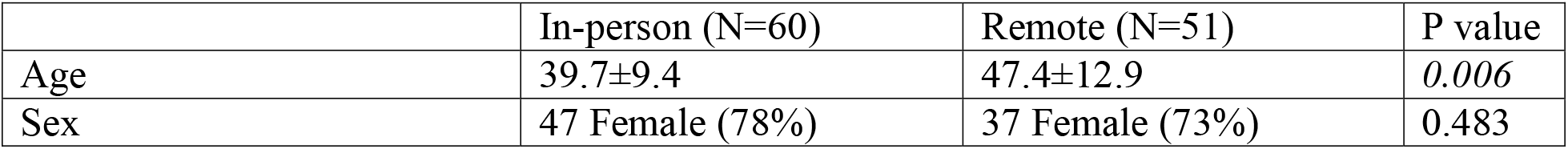
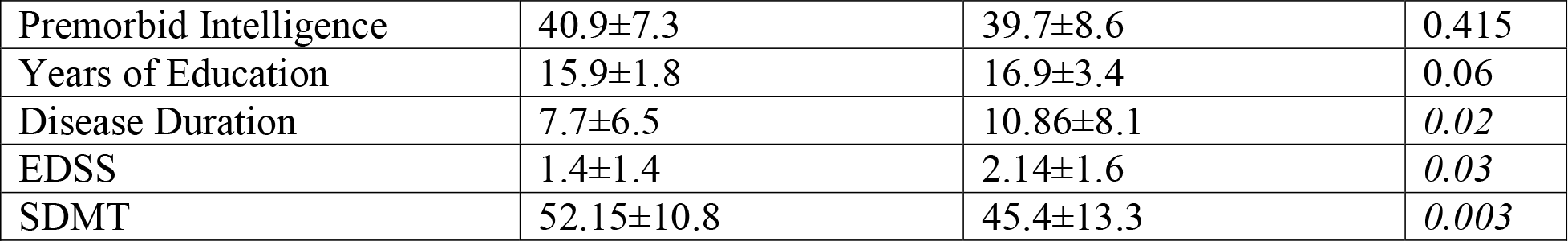
Demographic and clinical characteristics of the participants. Patients in the remote group were significantly older and had higher Expanded Disability Status Scale (EDSS) and lower Symbol Digit Modalities Test (SDMT) scores. There were no significant differences in other variables between groups. Premorbid verbal intelligence was estimated using the Wechsler Test of Adult Reading (WTAR). Variables were reported as mean ± standard deviation.

|  | In-person (N=60) | Remote (N=51) | P value |
| --- | --- | --- | --- |
| Age | 39.7±9.4 | 47.4±12.9 | *0.006* |
| Sex | 47 Female (78%) | 37 Female (73%) | 0.483 |
| Premorbid Intelligence | 40.9±7.3 | 39.7±8.6 | 0.415 |
| Years of Education | 15.9±1.8 | 16.9±3.4 | 0.06 |
| Disease Duration | 7.7±6.5 | 10.86±8.1 | 0.02 |
| EDSS | 1.4±1.4 | 2.14±1.6 | 0.03 |
| SDMT | 52.15±10.8 | 45.4±13.3 | 0.003 |

### 2.1. MRI

#### 2.1.1. MRI Data Collection

All MRI data were collected across a population of clinical scanners as part of routine evaluation at Columbia University Irving Medical Center.

For the patients in the remote group, 3 dimensional (3D) T1 (voxel resolution=0.5×0.5×0.6mm) and fluid-attenuated inversion recovery (FLAIR) images (voxel resolution=0.5×0.5×0.6mm) were extracted from the scans acquired during their clinical visits within +/- 1 year from the cognitive test administration (average time difference=95 days). MRI data were collected using 3 Tesla MRI scanners (GE Discovery, GE Healthcare, Wauwatosa, WI).

For the in-person group, there was no time gap between the day of SDMT administration and MRI scan as both were collected as part of the same research protocol. MRI data were collected using 3 Tesla MRI scanner (GE Discovery, GE Healthcare, Wauwatosa, WI). 3D T1 weighted images: Sagittal Structural T1 BRAVO sequence, voxel resolution =1×1×1mm and FLAIR images: voxel resolution =1×1×3mm.

#### 2.1.2. MRI Data Analysis

Total intracranial brain volume (ICV), total, cerebellar, cortical and deep gray matter volumes, also bilateral amygdala, caudate, nucleus accumbens, putamen, pallidus, hippocampus, and thalamus volumes separately, white matter hyperintensities, cerebral white matter volumes and brain parenchymal fraction were measured using FreeSurfer version 6.0,^27^ on 3D T1 images. In addition, volumes of 50 individual thalamic nuclei were calculated using the thalamic nuclei segmentation module.^28^ Since anterior thalamic nuclei was shown to be specifically involved in cognitive processes,^17,19^ from these 50 nuclei, we included only right and left anterior thalamic nucleus volumes in this study. Hemisphere-specific measures were averaged to obtain cross-hemispheric volumes, and ComBat harmonization was applied in order to control for MRI scanner and protocol differences between groups.^29^

Relevant relationships between brain measures and SDMT, age, sex, premorbid intelligence (IQ), and EDSS were preserved within the ComBat model.

### 2.2. Statistical analysis

To measure associations between MRI variables and SDMT scores, individual regression models were run for each MRI measure; SDMT was the outcome variable in each model, and all models controlled for patient age, sex, IQ, EDSS, and the difference (in days) between the measurement of SDMT and the relevant MRI scan. Models for whole tissue-class volumes (i.e., ICV, total cortical volume, total gray matter volume, and cerebral white matter volume) did not control for ICV, as its inclusion induces dependence across tissue classes; all other models included ICV as a covariate. To assess whether MRI-SDMT relationships differed between patients for whom SDMT was administered in person and those for whom it was administered remotely, an MRI measures-by-administration-technique interaction was included in each model.

In addition to testing for group-differences in each brain-SDMT relationship using interaction terms, we sought to assess the comparability of the two groups’ brain-SDMT effect sizes. First, we calculated the correlation between in-person effect sizes and remote effect sizes (across the 16 MRI measures tested); this measures the extent to which the strongest brain-SDMT relationships in one group also tend to be the strongest brain-SDMT relationships in the other. We also quantified the average difference between the in-person effect sizes and the remote effect sizes; this measured the extent to which the associations tend to be stronger on average in one group compared to the other. Confidence intervals for these values were calculated using 1000 bootstrap resamples. Finally, for each brain variable we quantified I^2^, a measure of heterogeneity that represents the percentage of variability in a coefficient that is attributable to groups (or studies, in the context of meta-analysis).

## 3. Results

In the primary analysis, we attempted replication of the well-validated relationship of lower SDMT score with reduced total gray matter volume that is a robust finding of MS studies. In these analyses, we have two sets of participants of similar size: (1) the “Remote” group (n=51) with SDMT obtained through video-conferencing as part of the MS Snapshot study’s protocol and volumetric measures derived from the clinical MRI closest to the video visit and (2) the “In-person” group (n=60) with SDMT and MRI obtained on the same day as part of the MemConnect study.

We note that the Remote participants were, on average, almost 8 years older, and, consistent with expected trends in MS disease progression, had a higher mean disability level (as measured by EDSS), longer disease duration and lower SDMT than the In-person group. The demographic and clinical characteristics of the groups are presented in **Table 1**.

We implemented our primary analysis as a meta-analysis between the two groups of participants with an interaction term for participant collection. Thus, we simultaneously assess the relation of SDMT with brain tissue volume measures and whether this relationship is different between the two sets of participants. As seen in **Table 2**, the replication meta-analysis for total gray matter volume is successful (p=0.005) and is in the expected direction given the positive correlation. However, the interaction term is not significant, reporting no meaningful difference between the two groups: the remote SDMT measure therefore captured the expected association of impaired cognition with brain atrophy.

**Table 2.**
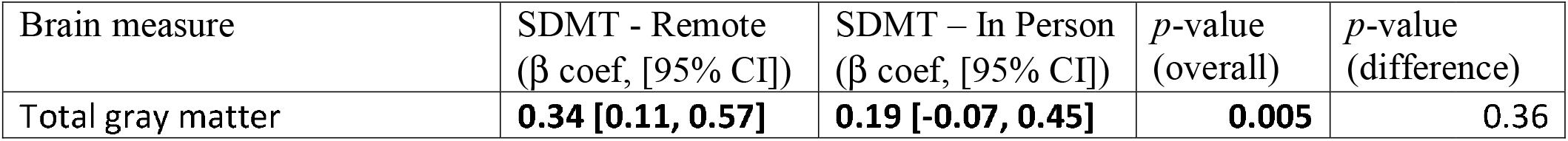
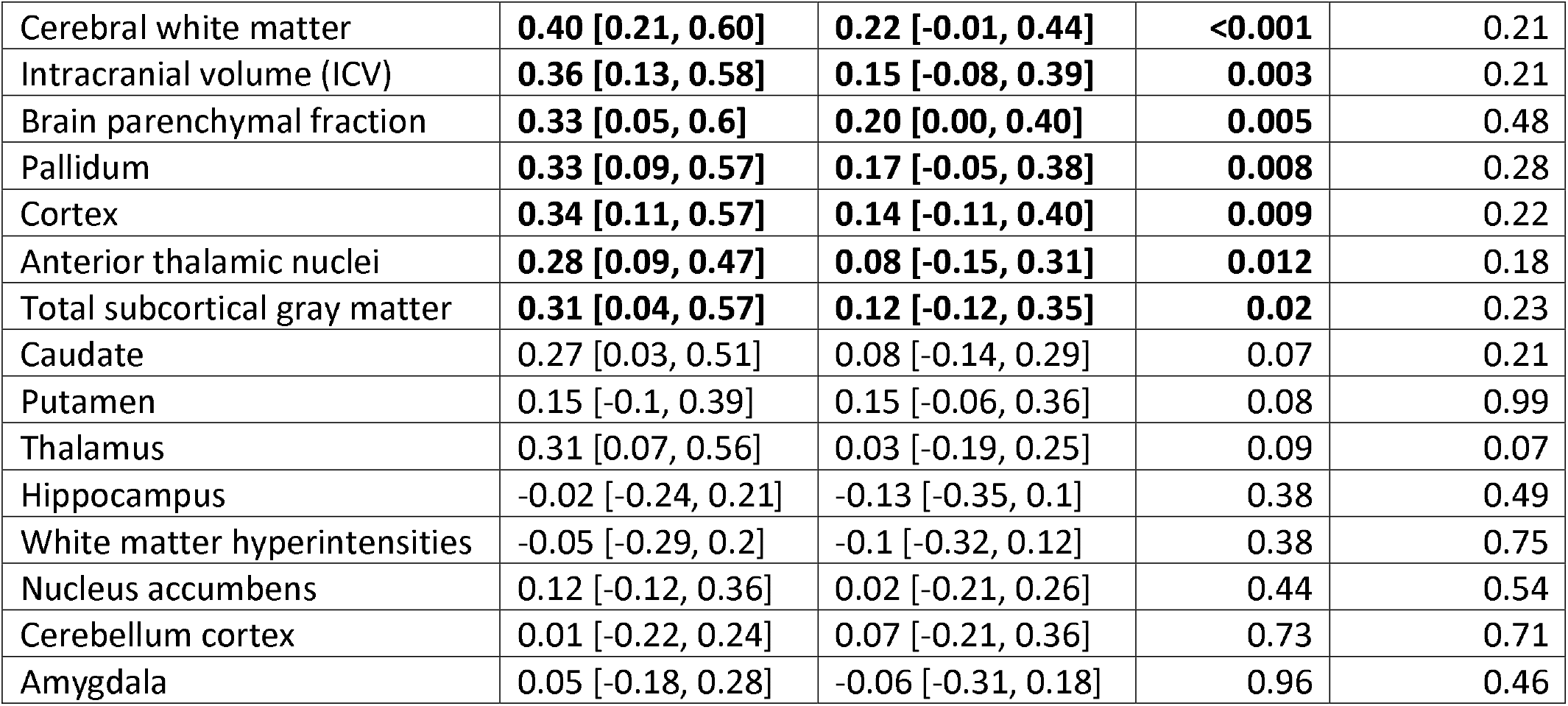
MRI-SDMT model results. For each of the 16 model MRI-SDMT relationships, columns represent the association within the remote administration group, the association within the in-person administration group, the p-value for the full-sample association, and the p-value for the interaction testing whether the association differs across groups. Associations are represented as standardized coefficients, and bolded associations were statistically significant in the full sample.

| Brain measure | SDMT - Remote<br>($\beta$ coef, [95% CI]) | SDMT – In Person<br>($\beta$ coef, [95% CI]) | *p*-value<br>(overall) | *p*-value<br>(difference) |
| --- | --- | --- | --- | --- |
| Total gray matter | **0.34 [0.11, 0.57]** | **0.19 [-0.07, 0.45]** | **0.005** | 0.36 |
| Cerebral white matter | **0.40 [0.21, 0.60]** | **0.22 [-0.01, 0.44]** | **<0.001** | 0.21 |
| Intracranial volume (ICV) | **0.36 [0.13, 0.58]** | **0.15 [-0.08, 0.39]** | **0.003** | 0.21 |
| Brain parenchymal fraction | **0.33 [0.05, 0.6]** | **0.20 [0.00, 0.40]** | **0.005** | 0.48 |
| Pallidum | **0.33 [0.09, 0.57]** | **0.17 [-0.05, 0.38]** | **0.008** | 0.28 |
| Cortex | **0.34 [0.11, 0.57]** | **0.14 [-0.11, 0.40]** | **0.009** | 0.22 |
| Anterior thalamic nuclei | **0.28 [0.09, 0.47]** | **0.08 [-0.15, 0.31]** | **0.012** | 0.18 |
| Total subcortical gray matter | **0.31 [0.04, 0.57]** | **0.12 [-0.12, 0.35]** | **0.02** | 0.23 |
| Caudate | 0.27 [0.03, 0.51] | 0.08 [-0.14, 0.29] | 0.07 | 0.21 |
| Putamen | 0.15 [-0.1, 0.39] | 0.15 [-0.06, 0.36] | 0.08 | 0.99 |
| Thalamus | 0.31 [0.07, 0.56] | 0.03 [-0.19, 0.25] | 0.09 | 0.07 |
| Hippocampus | -0.02 [-0.24, 0.21] | -0.13 [-0.35, 0.1] | 0.38 | 0.49 |
| White matter hyperintensities | -0.05 [-0.29, 0.2] | -0.1 [-0.32, 0.12] | 0.38 | 0.75 |
| Nucleus accumbens | 0.12 [-0.12, 0.36] | 0.02 [-0.21, 0.26] | 0.44 | 0.54 |
| Cerebellum cortex | 0.01 [-0.22, 0.24] | 0.07 [-0.21, 0.36] | 0.73 | 0.71 |
| Amygdala | 0.05 [-0.18, 0.28] | -0.06 [-0.31, 0.18] | 0.96 | 0.46 |

We then extended our evaluation with a set of secondary analyses that evaluated the relationship of SDMT to 16 other MRI-derived volumetric measures (**Table 2**). Seven of these measures also had a nominal (p<0.05) association with SDMT in the meta-analysis: cerebral white matter, intracranial, cerebral, cerebellar gray matter, pulvinar and anterior thalamic nuclei volumes and brain parenchymal fraction. Allowing the MRI-SDMT association to vary as a function of SDMT administration modality did not reveal any significant interaction effects across the 16 models, such that administration modality did not appear to meaningfully affect MRI-SDMT association strength (interaction p values ranged from 0.07 and 0.99, see **Figure 1a**).

**Figure 1.**
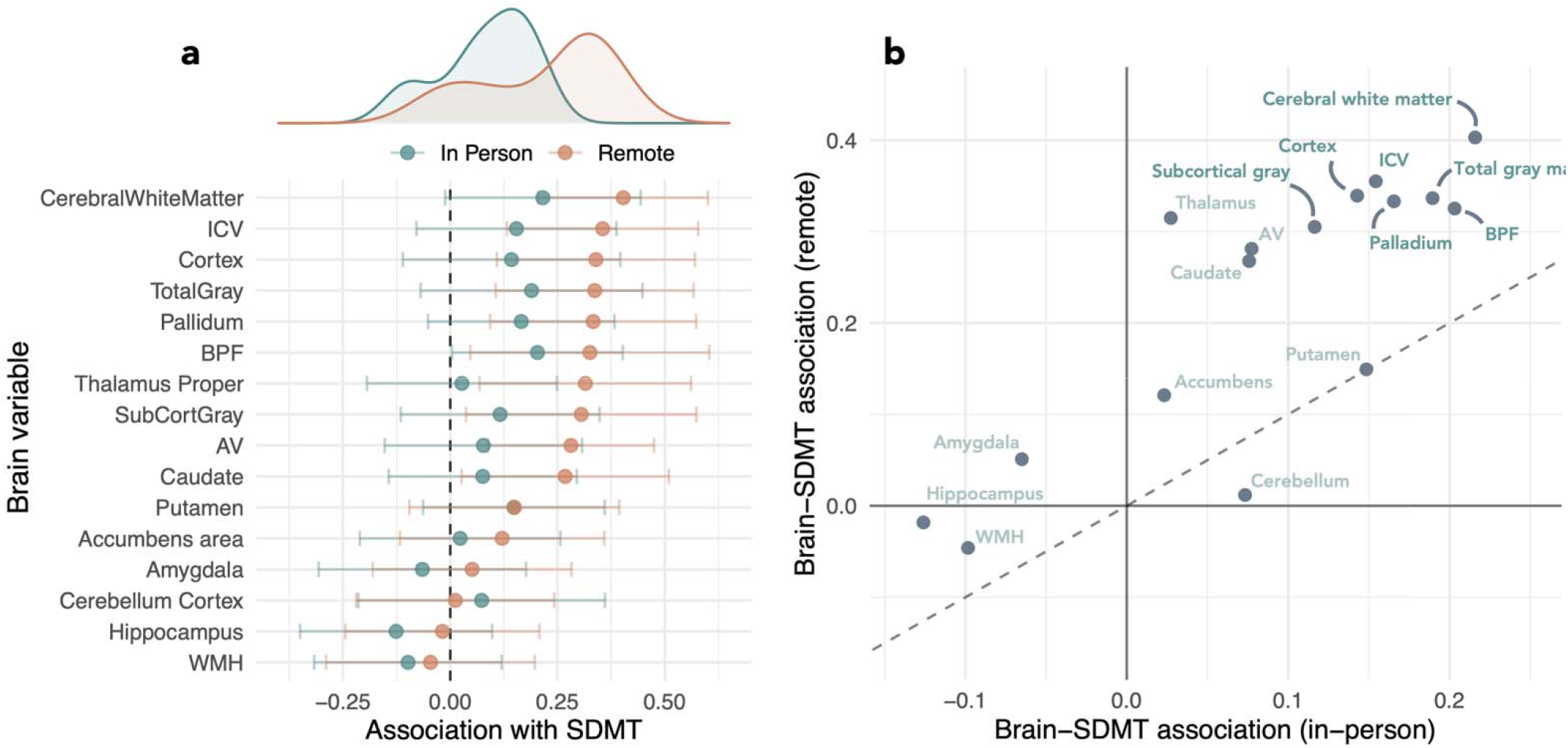
**(a)** Distribution of brain-SDMT effect sizes across regions and across SDMT administration types. Top density plots show the overall distribution, dot-and-whiskers show the region-specific effect sizes and uncertainty. **(b)** Scatterplot comparing brain-SDMT effect sizes in the in-person SDMT group (x-axis) to those in the remote SDMT group (y-axis). The dotted line represents y=x correspondence. Regions with strong SDMT associations in both groups are labeled in bold. Abbreviations: ICV=Intracranial volume, AV=Anterior thalamic nuclei, BPF= Brain parenchymal fraction, WMH= White matter hyperintensities, SDMT=Symbol Digit Modalities Test.

The correspondence between each group’s MRI-SDMT associations was measured by calculating the correlation across regions, indicating the extent to which regions with high associations in one group tended to also show high associations in the other. These values were significantly correlated suggesting good correspondence (*r* = 0.83; 95% CI = [0.07, 0.86]; see Figure 1b). We additionally assessed whether the remote assessment group’s r-values tended to be lower or higher than those in the in-person group by calculating the average difference between the measures across brain regions. Although correlations were stronger in the Remote group, the difference was not significant (β_Remote_ - β_In-person_ = 0.14, 95% CI = [-0.04, 0.34]).

Across MRI measures, the average I^2^ value was 15%, reflecting very little heterogeneity between the two sets of participants. Eleven of the 16 MRI measures showed I^2^ values less than 25% (generally considered small), and 5 more showed values less than 50% (generally considered medium). Only the thalamus showed a medium-to-large I^2^ of 66%, on this measure the two sets of participants may demonstrate some heterogeneity (i.e., a proportionally greater loss of thalamic volume in the Remote group).

## 4. Discussion

Previous studies have demonstrated the reliability of remotely administered cognitive batteries in MS as a part of telehealth assessments.^7^ Here, we took the next step and evaluated the practical utility of these telehealth assessments to power clinical research studies. We chose one of the more robust associations in the MS literature to drive this study, and therefore evaluated the relation of SDMT to brain atrophy.

Our results showed that the mode of SDMT administration did not meaningfully affect the association of SDMT with brain tissue volumes: we replicated the expected results that have been shown in a variety of patient collections. Given our moderate sample size and baseline differences between the Remote and In-person groups, we cannot exclude the possibility that there may be a small difference in the magnitude of the SDMT-brain volume relationship between the two approaches; however, this would not affect the fundamental result that an SDMT obtained through video-conferencing provides similar results to that obtained in person. We thus extend the prior results of good correspondence between Remote and In-person testing in the same individuals^7,9^ to a successful field test of a video-conference based cognitive measure.

*A priori*, one might have expected the effect size of the associations to be more modest and less significant in the Remote group both because the remote SDMT may be less accurate than the in-person variety and because the Remote group was hobbled with the use of repurposed clinical MRI instead of the dedicated MRI collected in the MemConnect study (on the same day as the in-person SDMT). However, the MRI-SDMT associations were stronger in the Remote group, although this difference was not significant. This difference is most likely due to the fact that the Remote group was older, had higher disease burden and longer disease duration (overall greater variance in the measures under study) and not any advantage of the video-conference measure. We found that brain tissue volumes showing high associations with the SDMT in one group also showed high associations in the other (*r* = 0.83; 95% CI = [0.07, 0.86]) which means, there was a consistent MRI measure-SDMT performance association pattern between two groups. The brain regions found to be correlated with SDMT in our study are in line with the findings of previous studies investigating brain tissue volume-cognition association in MS.^30^ However, it is important to note that, although thalamus volume has often been emphasized as an important predictor of cognition in MS,^17,19,20^ in our study it showed a medium-to-large I^2^ (66%), which may demonstrate some heterogeneity between the groups. Indeed, it was significantly associated with the SDMT only in the remote patient group who are clinically more advanced (*r* = 0.32; 95% CI = [0.07, 0.57]).

Another aspect of this study was leveraging MRI data which were originally collected for clinical purposes. Using T1 images, we extracted a comprehensive collection of gray and white matter metrics which were shown to be correlated with SDMT performance in the previous studies. However, future studies using dedicated research imaging should investigate the associations between other MRI measures (e.g., white matter microstructural integrity, lesion load/type/location, functional connectivity) and test administration modalities as well.

Telehealth applications enabled patients with reduced mobility and/or geographical constraints to access specialists and improved patient and caregiver convenience, comfort and safety constraints (factors which became particularly important during the global pandemic) over the last couple of years.^31–33^ While supporting their use, the American Academy of Neurology (AAN) also highlighted important research and practical knowledge gaps in these applications (e.g., lack of normative data and standardized administration techniques, variability in insurance coverage policies, socio-economic and technical difficulties like having privacy and confidentiality, a proper computer/monitor and fast internet connection at home, age and education related barriers to technology use, etc.).^34^ Such concerns have been raised by the researchers studying telehealth applications in MS as well.^35,36^ Therefore, future studies should address these issues and eventually a consensus on a comprehensive remote cognitive testing battery (including other frequently used tests in cognitive monitoring in MS) with standardized administration techniques and normative data should be established.

Our study has limitations. First, MRI scanners and image acquisition parameters were not the same for the two patient groups that we compared. To mitigate the effects of scanner differences, we used a well-established statistical method/tool (i.e., ComBat) to control the variability due to scanner/protocol differences, although it must be acknowledged that scanner differences may still exert an effect on our results. Secondly, there was no time difference between the MRI scan and cognitive testing date in the in-person group, however there was a time gap for the remote group. Although we limited this gap to +/- 1 year and defined this as a covariate and controlled for it in our analyses, the MRI-SDMT associations in the remote group may not perfectly reflect the up-to-date relationship between the brain tissue volume and cognitive performance.

## 5. Conclusion

In conclusion, our study showed consistent relationships to neural correlates across in-person and remote SDMT administration modalities, extending the findings of the previous studies demonstrating the feasibility of remote administration of the SDMT. Most of these studies were conducted by administering SDMT to the same participants both in-person and remotely. To our knowledge, this is the first study investigating the relationship of remote and in-person administration modalities in two independent patient groups to MRI measures as an external outcome. Hence, our study can be considered a step towards providing predictive validity to remote administration of the SDMT in MS.

## Data Availability

All data produced in the present study are available upon reasonable request to the authors

## Author Statement

Korhan Buyukturkoglu: Conceptualization, methodology, MRI data Analysis, writing.

Jordan Dworkin: Methodology, statistical analysis, writing and editing.

Victor Levia: MRI data Analysis.

Frank Provenzano: Conceptualization, methodology, editing– original draft, MRI data Analysis.

Pamela Guevara: Methodology.

Phil De Jager, Victoria Leavitt, Claire Riley: Conceptualization, methodology, editing– original draft, supervision.

